# Perceived Stress During Gender-Affirming Hormone Therapy: A Comparative Study of Feminizing and Masculinizing Hormone Therapy in an International Multicenter Prospective Cohort

**DOI:** 10.1101/2025.05.01.25326821

**Authors:** Marijn Schipper, Margot W. L. Morssinkhof, Baudewijntje P. C. Kreukels, Guy T’Sjoen, Alessandra D. Fisher, Yona Greenman, Karin van der Tuuk, Martin den Heijer, David Matthew Doyle, Birit F. P. Broekman

## Abstract

Cisgender women report higher stress than cisgender men, potentially due to psychosocial and biological factors, including sex hormone levels. Gender-affirming hormone therapy (GAHT) alters hormone levels, but its impact on perceived stress remains unclear. This study examined changes in perceived stress after 3 and 12 months of GAHT and potential differences between feminizing (FHT) and masculinizing hormone therapy (MHT).

Data were drawn from two prospective cohort studies (ENIGI and RESTED) in the Netherlands, Belgium, and Israel. A total of 442 individuals (median age 23 years, IQR 20.5 to 28.0) completed the 10-item Perceived Stress Scale (PSS) before starting GAHT and after 3 and 12 months. Linear mixed models assessed changes after starting GAHT and differences between FHT and MHT groups.

Baseline perceived stress levels did not significantly differ between groups (0.15, p = 0.84). No significant changes in perceived stress were observed after 3 or 12 months, nor were there significant differences in changes between FHT and MHT (-1.0, p = 0.21; 0.25, p = 0.76).

Improvements in well-being during GAHT may not reduce perceived stress, potentially due to ongoing gender minority stressors. Future research should explore stressors and coping mechanisms to identify strategies for reducing perceived stress during GAHT.

## Introduction

Stress is a significant issue in modern society; the American Psychological Association (2022) reported that 76% of adults experienced stress in the past month, negatively impacting their physical and mental health. Although moderate, short-term stress can be beneficial, excessive or prolonged stress impairs daily functioning and increases the risk of physical and mental health disorders (Gao et al., 2022; Russell & Lightman, 2019).

Cortisol indicates a physiological stress response, whereas self-reported perceived stress is a key clinical parameter influencing overall health (Keller et al., 2012). Both are influenced by external stressors and coping resources, but perceived stress specifically reflects an individual’s subjective appraisal of stress (Folkman, 2020). Studies found that cisgender women report higher perceived stress than cisgender men on average (Graves et al., 2021; Kelly et al., 2008; Thorsén et al., 2022; Yalcin-Siedentopf et al., 2021). The biopsychosocial model may explain gender differences as a result of the interplay between social, psychological, and biological factors (Bennett et al., 2018). Socially, women face greater exposure to gender-based violence, discrimination, and economic inequalities, which can contribute to higher stress (Graves et al., 2021; Kuehner, 2017; Riecher-Rössler, 2017; Seedat et al., 2009). Psychologically, differences in emotional regulation, personality traits, and prior experiences may influence stress perception and coping mechanisms (Salk et al., 2017; Stoica et al., 2021). Biologically, sex hormones influence the hypothalamic-pituitary-adrenal (HPA) axis, impacting cortisol levels. Variations in HPA axis reactivity have been observed between men and women, particularly during the follicular phase or in women using oral contraceptives (Henze et al., 2021; Stephens et al., 2016), as well as in women across the menstrual cycle, puberty, and menopause (Gordon et al., 2016; Hoyt & Falconi, 2015; Kirschbaum et al., 1999; Leistner & Menke, 2020; Smith, 2021; Woods et al., 2006), highlighting the role of sex hormones in stress regulation. While the influence of sex hormones on cortisol is well-documented, it remains unclear whether changes in sex hormone levels directly affect perceived stress.

Transgender individuals experience more psychosocial stress and lower quality of life than cisgender individuals (Collet et al., 2023; Doyle et al., 2023; Mezza et al., 2024). These disparities are largely attributed to gender minority stress, which includes discrimination, stigma, and misgendering (Mezza et al., 2024; Singh et al., 2023). Additionally, transgender individuals tend to have smaller social networks, further impacting stress perception and limit coping resources (Collet et al., 2023).

Gender-affirming hormone therapy (GAHT) has been shown to reduce psychological distress and improve quality of life (Baker et al., 2021; Chen et al., 2023; Colizzi et al., 2013; Doyle et al., 2023; van Leerdam et al., 2023). Colizzi et al. (2013) reported a significant decrease in perceived stress after 12 months of GAHT, with no differences between feminizing hormone therapy (FHT) and masculinizing hormone therapy (MHT). However, their study had limitations, such as a small sample size and potential access-to-care biases due to participants awaiting hormone therapy approval at baseline (Call et al., 2021; Turban et al., 2022).

It remains unclear whether FHT and MHT differentially affect perceived stress. Biologically, FHT has been associated with greater fluctuations in negative affect compared to MHT (Morssinkhof et al., 2025). This may contribute to heightened stress perceptions, as increased emotional variability is linked to lower psychological well-being (Houben et al., 2015). Additionally, 12 months of FHT has been associated with increased low mood whereas this was not found after MHT (Morssinkhof et al., 2024). Psychosocially, individuals initiating FHT may experience more gender minority stress than those starting MHT, potentially influencing stress outcomes (Poquiz et al., 2021).

This study aimed to examine changes in perceived stress after 3 and 12 months of GAHT and to explore potential differences between FHT and MHT. Based on previous research (Colizzi et al., 2013), we expected both groups to report reduced stress levels over time. However, we hypothesized that this reduction would be less pronounced among individuals starting FHT due to an increase in negative affect variability after starting GAHT (Morssinkhof et al., 2025) and higher levels of gender minority stress (Poquiz et al., 2021).

## Methods

### Study setting

This study used data from two multicenter prospective cohort studies: the ENIGI study, investigating the effects of GAHT (Dekker et al., 2016), and the RESTED study, examining sleep and mood changes following GAHT (Morssinkhof et al., 2024). The ENIGI study involved centers in Belgium, the Netherlands, Italy, Norway, and Israel (Dekker et al., 2016); however, only Amsterdam University Medical Center (the Netherlands), Ghent University Hospital (Belgium), and Tel Aviv Sourasky Medical Center (Israel) administered the questionnaire used in the current study. Baseline measurements were collected from December 2019 to January 2024. The RESTED study recruited participants from Amsterdam University Medical Center and University Medical Center Groningen (the Netherlands), with baseline measurements between December 2019 and August 2022. Ethical approval was obtained (ENIGI: 2019.469, RESTED: 2019.353), and all participants provided informed consent.

### Participants

This secondary analysis included individuals initiating GAHT at a collaborating institute who could speak, read, and write the local language. No power analysis was conducted a priori, as pre-existing databases with fixed samples were utilized. RESTED participants were aged between 18 and 50, while ENIGI participants were over 17 in Ghent and over 18 in Amsterdam and Tel Aviv. Additional exclusion criteria of the RESTED study included a diagnosed sleep disorder or benzodiazepine/opiate use. Participants needed to have completed the 10-item Perceived Stress Scale (PSS) at least once (baseline, 3 months, and/or 12 months). Exclusion criteria were having used GAHT before or using medications severely affecting sex hormones (e.g., tamoxifen). Menstrual cycle regulation alongside MHT was allowed, and we conducted a sensitivity analyses adjusting for use of menstrual cycle regulation. For participants enrolled in both studies, duplicate measurements were removed, with priority given to RESTED data due to its more detailed demographic information (Figure 1).

**Figure 1.**
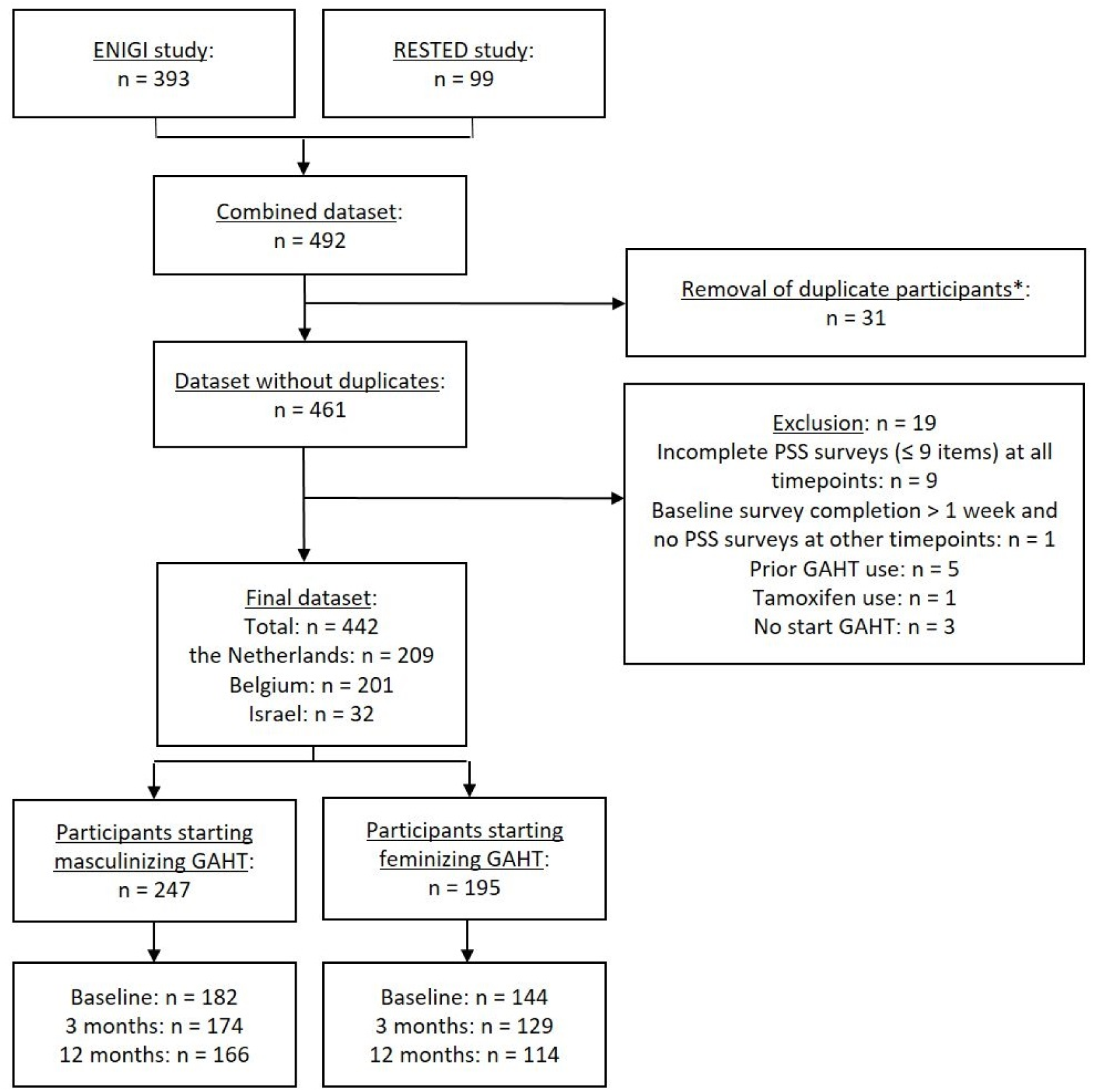
Flow chart showing sample sizes at inclusion and follow-up. *Duplicate measurements from participants enrolled in both studies were excluded from the dataset.

### Procedure

Participants completed online questionnaires at baseline and after 3 and 12 months of GAHT, corresponding to the timing of clinic visits. ENIGI baseline questionnaires were administered at GAHT initiation, whereas RESTED participants completed them weeks or months before starting GAHT. ENIGI questionnaires which were completed over a week after starting GAHT were removed. Clinical data (sex assigned at birth, age, substance use, GAHT type, cycle regulation, and psychotropic medication use) were extracted from medical records. Missing psychotropic medication data were carried forward from earlier time points to include as a covariate in analyses. Blood levels of testosterone and estradiol were assessed at baseline, and after 3 and 12 months of GAHT (Supplementary materials 1), primarily for therapy adherence monitoring rather than causal inference.

### Gender-affirming hormone therapy

MHT included testosterone administered via gel (50 mg daily) or intramuscular injections (esters: 250 mg/2 weeks; undecanoate: 1000 mg/12 weeks). FHT included estradiol, used orally (2 mg twice daily) or transdermally (patch: 100 µg/24h twice weekly; spray: 4.59 mg daily; gel: 1.5 mg daily) with anti-androgens (cyproterone: 10-50 mg daily; spironolactone: 25-100 mg daily) or GnRH analogues (leuprorelin: 3.75 mg/4 weeks; triptorelin: 3.75 mg/4 weeks or 11.25 mg/12 weeks).

### Perceived stress

Perceived stress was measured using the validated 10-item PSS (Cohen et al., 1983), a widely used self-report scale with good psychometric properties, including high internal consistency (Cronbach’s α = 0.74-0.91 across different populations) (Lee, 2012; Yılmaz Koğar & Koğar, 2024). The PSS assesses stress experienced in the past month through items such as: “How often have you been upset because of something that happened unexpectedly?” and “How often have you felt difficulties were piling up so high that you could not overcome them?” (Cohen et al., 1983). Responses were rated on a 5-point scale (0 = never, 4 = very often). Items 4, 5, 7, and 8 were reverse-coded, and total scores (0-40) were calculated by summing all item scores, with higher values indicating greater stress.

### Statistical analyses

Statistical analyses were performed in RStudio (R Development Core Team, 2010) using lmerTest (Kuznetsova et al., 2017). PSS data were included if at least 9 of 10 items were completed; mean imputation was applied for one missing item. Continuous variables were summarized using means (SD) or medians (IQR) depending on normality distribution. Results were reported as effect estimates with 95% CIs and p-values (<0.05 considered significant).

Linear mixed models assessed baseline PSS scores and changes after starting GAHT, with time as a fixed predictor (i.e., comparison between baseline and 3 and 12 months of GAHT) and random intercepts for participants nested within countries to model both individual- and country-level effects on PSS scores, accounting for social context variability (e.g., average levels of gender minority stress exposure which could differ between countries) (Moerbeek, 2004; Snijders & Bosker, 1999). This structure better fit the data than treating country as a covariate (Supplementary Materials 2).

Stratified analyses examined changes separately per GAHT type (MHT/FHT), and interaction terms tested between-group differences at baseline, 3, and 12 months. Models were adjusted for age and psychotropic medication use.

Sensitivity analyses (Supplementary materials 3–5) involved excluding participants with data from only one or two time points, excluding those receiving FHT without anti-androgens, and adjusting for cycle regulation in MHT users.

## Results

### Participants

The study included 492 participants: 393 from the ENIGI study and 99 from the RESTED study. A total of 31 participants participated in both studies, and duplicate measurements of these participants were removed, prioritizing RESTED data. In total, 52 individual questionnaires were removed: 26 (6%) incomplete questionnaires (≤9 items), and 26 (7%) baseline questionnaires which were completed >1 week after GAHT initiation. In total, 19 (4%) participants were excluded from the database. 9 (2%) participants were excluded since they had not completed a questionnaire at any time point, and only one participant (0.2%) was excluded because they completed the baseline questionnaire >1 week after GAHT initiation and did not contribute any data at any other timepoint. Further exclusions included prior GAHT use (n = 5, 1%), tamoxifen use (n = 1, 0.2%), and not initiating GAHT (n = 3, 1%). The final dataset included 442 participants: 209 (47%) from the Netherlands, 201 (45%) from Belgium, and 32 (8%) from Israel. Of these, 247 (56%) started MHT and 195 (44%) FHT. Baseline PSS data were available for 182 (74%) MHT and 144 (74%) FHT participants, with 154 (35%) providing data at all time points. Missing data resulted from loss to follow-up as well as from pending study invitations, as the ENIGI study is ongoing. Figure 1 illustrates the sample sizes.

### Demographic characteristics

Participants’ median age was 23.0 years (IQR 20.5 to 28.0). Psychotropic medication was used by 78 (18%) participants, and 36 (19%) of those starting MHT used cycle regulation. Six (3%) participants received estrogens without anti-androgens. Table 1 presents demographics.

**Table 1.** Demographic characteristics of the study participants.

| **Table 1.** Demographic characteristics of the study participants |  |  |  |
| --- | --- | --- | --- |
|  |  | **Participants starting MHT** | **Participants starting FHT** |
| **Sample size (n, % of total study sample)** |  | 247 (56%) | 195 (44%) |
| **Age (median, IQR)** |  | 22.0 (20.1 to 25.3) | 25.2 (21.7 to 30.0) |
| **Country (n, %)** | *The Netherlands* | 114 (46%) | 95 (49%) |
|  | *Belgium* | 124 (50%) | 77 (39%) |
|  | *Israel* | 9 (4%) | 23 (12%) |
| **Alcohol use (median, IQR)** | *Consumptions per week* | 0 (0.0 to 2.0) | 0.5 (0.0 to 2.0) |
|  | *Missing* | 24 (10%) | 22 (11%) |
| **Smoking (n, %)** | *No smoking* | 169 (68%) | 146 (76%) |
|  | *Smoking* | 54 (22%) | 27 (14%) |
|  | *Missing* | 24 (10%) | 22 (11%) |
| **Drug use (n, %)** | *No drug use* | 191 (77%) | 155 (79%) |
|  | *Drug use* | 32 (13%) | 23 (12%) |
|  | *Missing* | 24 (10%) | 17 (9%) |
| **Psychotropic medication use (n, %)** | *No psychotropic medication* | 184 (74%) | 150 (77%) |
|  | *Psychotropic medication* | 48 (19%) | 30 (15%) |
|  | *Antidepressants* | 38 (15%) | 20 (10%) |
|  | *Stimulants* | 14 (6%) | 12 (6%) |
|  | *Antipsychotics* | 13 (5%) | 9 (5%) |
|  | *Anxiolytics* | 5 (2%) | 0 |
|  | *Mood stabilizers* | 2 (1%) | 0 |
|  | *Missing* | 15 (6%) | 15 (8%) |
| **Cycle regulation use at baseline (n, %)** | *No cycle regulation* | 141 (77%) | - |
|  | *Cycle regulation* | 36 (19%) | - |
|  | *Missing* | 5 (3%) | - |
| **Form of testosterone at start of GAHT (n, %)** | *Testosterone gel* | 100 (40%) | - |
|  | *Long-acting injections* | 87 (35%) | - |
|  | *Short-acting injections* | 59 (24%) | - |
|  | *Oral testosterone* | 1 (0%) | - |
| **Form of estrogen at start of GAHT (n, %)** | *Estradiol tablets* | - | 136 (70%) |
|  | *Estradiol patches* | - | 41 (21%) |
|  | *Estradiol gel* | - | 16 (8%) |
|  | *Estradiol spray* | - | 2 (1%) |
|  | *None\** | - | 0 |
| **Form of anti-androgen at start of GAHT (n, %)** | *GnRH analogues (triptoreline, leuproreline)* | - | 106 (54%) |
|  | *Cyproterone acetate* | - | 75 (38%) |
|  | *Spironolactone* | - | 8 (4%) |
|  | *None\** | - | 6 (3%) |
| * Six participants received estrogens without anti-androgens; sensitivity analysis excluding them is in Supplementary Materials 4. |  |  |  |

### Perceived stress before starting GAHT

Baseline PSS scores averaged 17.1 (SD 7.2) in the MHT group and 16.3 (SD 7.2) in the FHT group, with no significant difference between groups (after nesting and adjusting for covariates: b = 0.15, 95% CI -1.4 to 1.7, p = 0.84). Table 2 provides full baseline PSS data.

**Table 2.** Perceived stress outcomes before starting GAHT (baseline)

| **Table 2.** Perceived stress outcomes before starting GAHT (baseline) |  |  |  |
| --- | --- | --- | --- |
|  |  | **Perceived stress – raw model\*** | **Perceived stress – adjusted model\*\*** |
| **FHT group vs. MHT group** | *Estimate (95% CI), p* | *-0.90 (-2.4 to 0.7)*<br>*P = 0.26* | *0.15 (-1.4 to 1.7)*<br>*P = 0.84* |
| *Participants nested within countries (i.e., the Netherlands, Belgium or Israel). |  |  |  |
| **Participants nested within countries, and adjusted for age and psychotropic medication use. |  |  |  |

### Perceived stress following 3 and 12 months of GAHT

No significant changes in PSS scores occurred after GAHT initiation (3 months: b = -0.08, 95% CI -0.7 to 0.9, p = 0.83; 12 months: b = 0.61, 95% CI -0.2 to 1.4, p = 0.14). Interaction analyses showed no significant differences between MHT and FHT over time (3 months: b = -1.0, 95% CI -2.6 to 0.6, p = 0.21; 12 months: b = 0.25, 95% CI -1.4 to 1.9, p = 0.76). Table 3, Table 4 and Figure 2 summarize these results.

**Figure 2.**
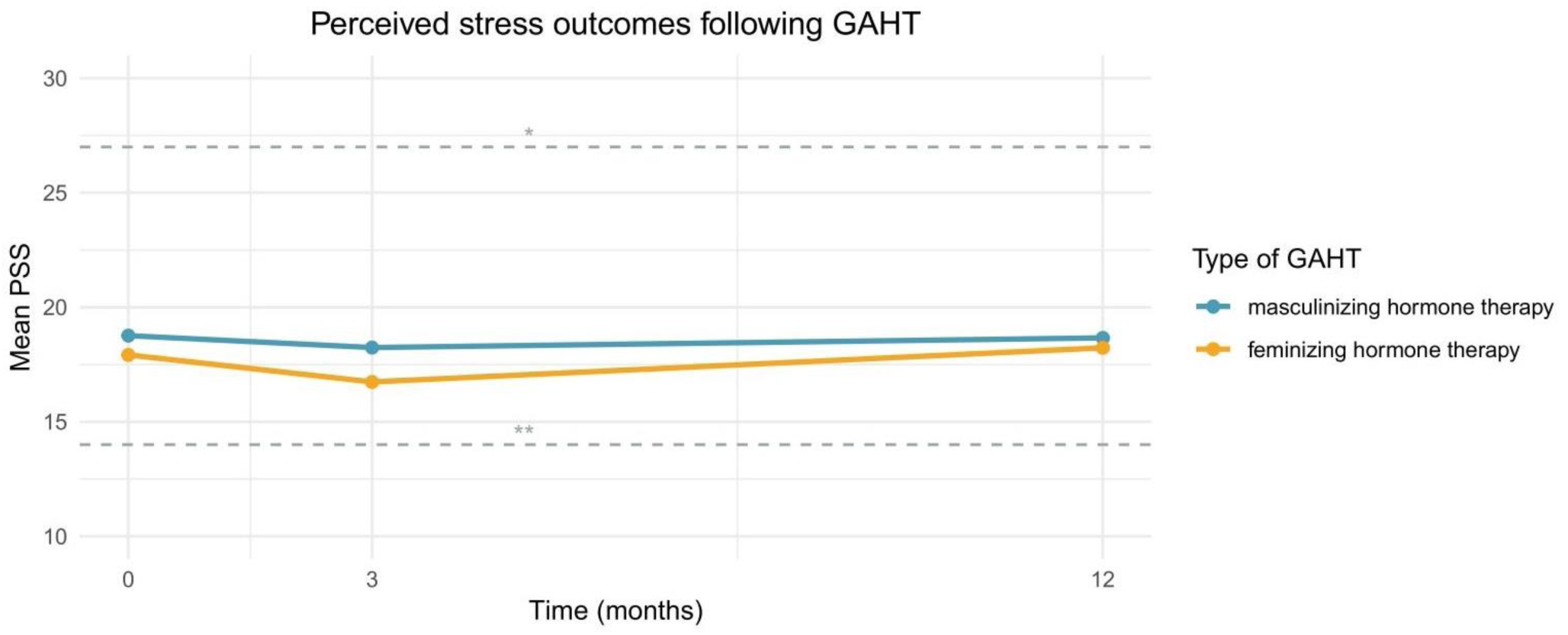
Mean PSS scores by group before GAHT and after 3 and 12 months. Blue: MHT, orange: FHT. Horizontal lines represent stress severity categories: the region between the dashed lines corresponds to moderate perceived stress (14-26).

**Table 3.**
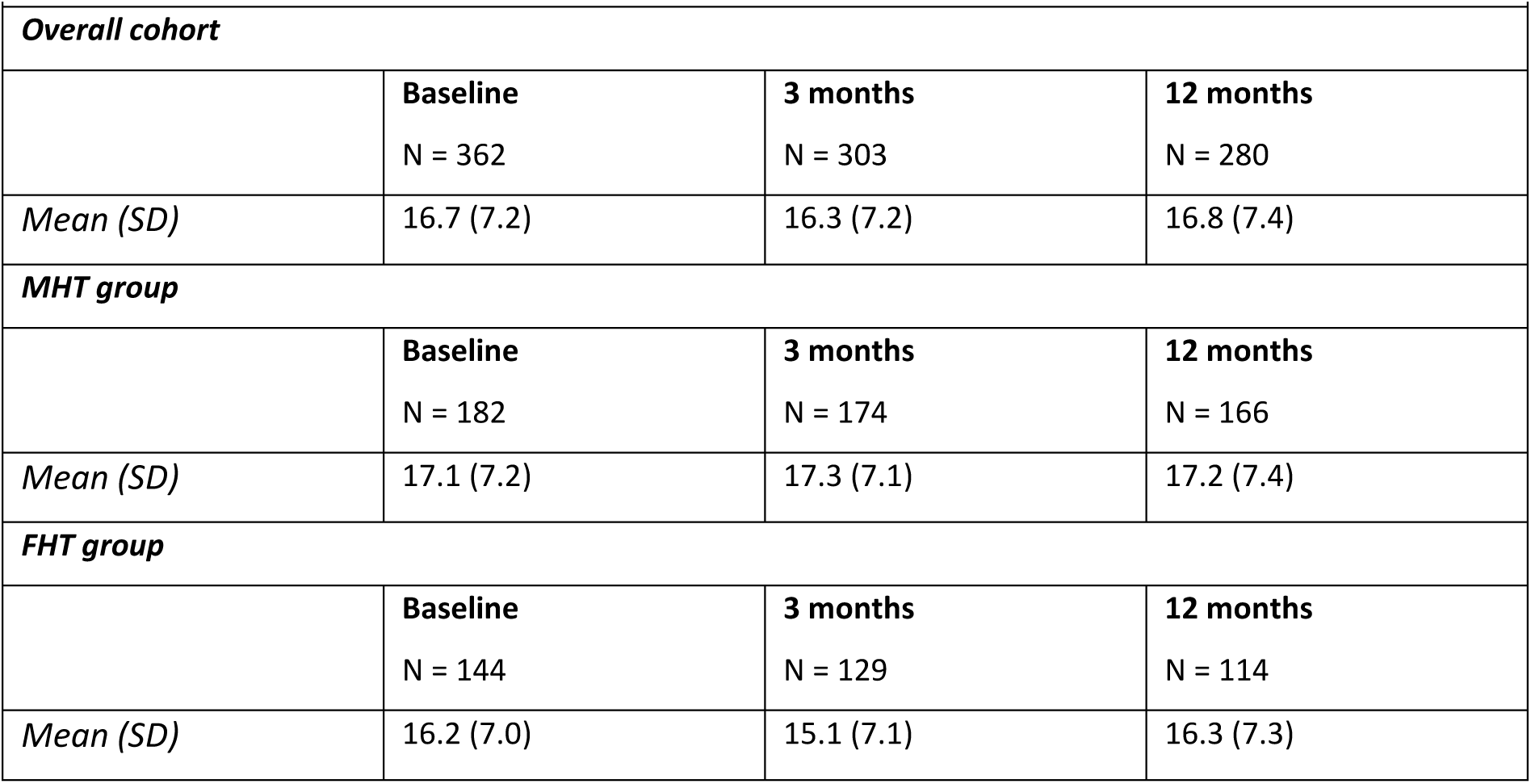
Mean perceived stress scores at baseline and after 3 and 12 months of GAHT.

**Table 4.**
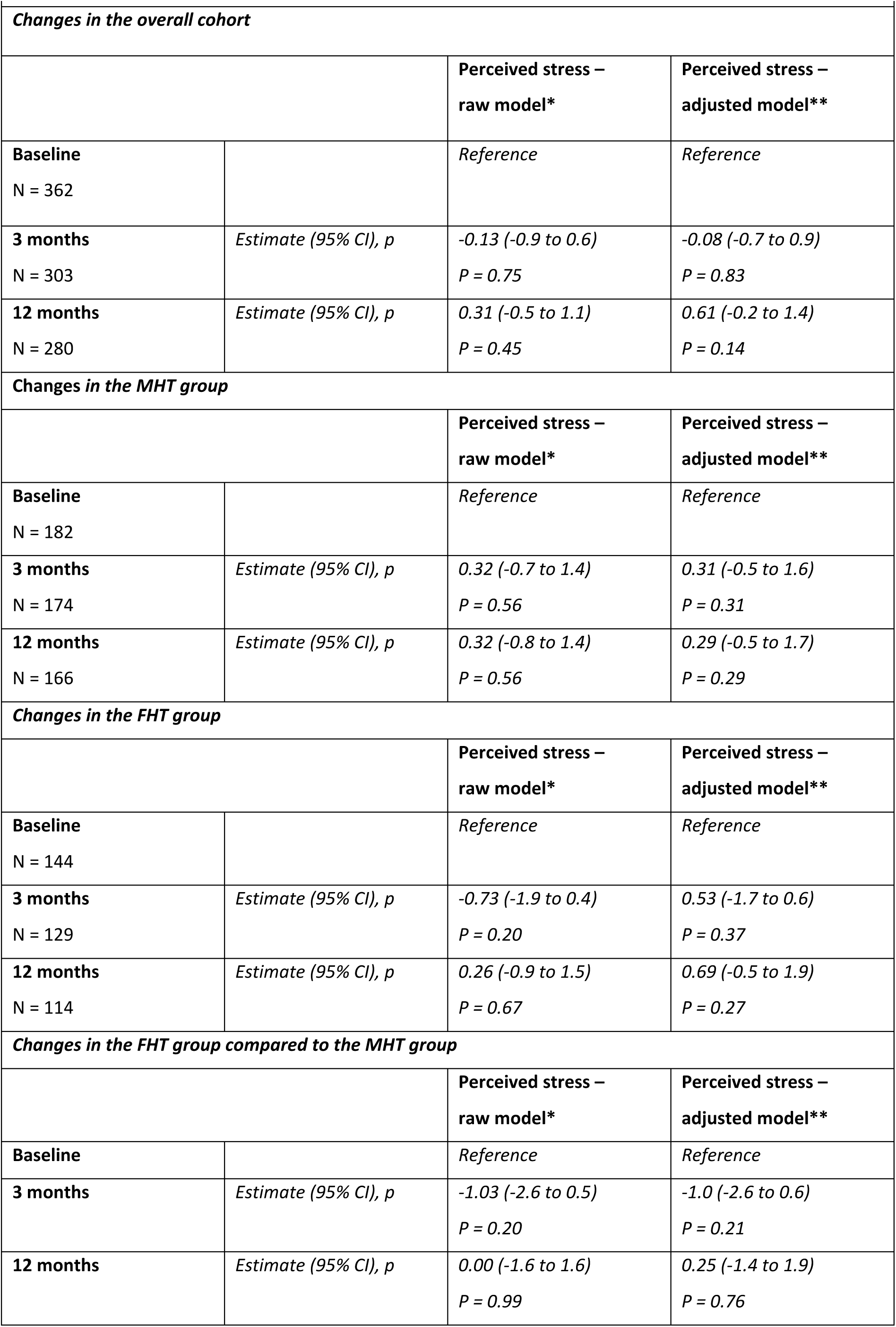
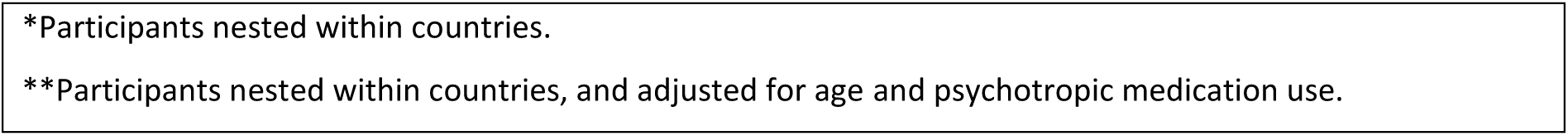
Perceived stress changes after GAHT.

| **Table 4.** Perceived stress changes after GAHT |  |  |  |
| --- | --- | --- | --- |
| *Changes in the overall cohort* |  |  |  |
|  |  | **Perceived stress –**<br>**raw model\*** | **Perceived stress –**<br>**adjusted model\*\*** |
| **Baseline**<br>N = 362 |  | *Reference* | *Reference* |
| **3 months**<br>N = 303 | *Estimate (95% CI), p* | *-0.13 (-0.9 to 0.6)*<br>*P = 0.75* | *-0.08 (-0.7 to 0.9)*<br>*P = 0.83* |
| **12 months**<br>N = 280 | *Estimate (95% CI), p* | *0.31 (-0.5 to 1.1)*<br>*P = 0.45* | *0.61 (-0.2 to 1.4)*<br>*P = 0.14* |
| *Changes in the MHT group* |  |  |  |
|  |  | **Perceived stress –**<br>**raw model\*** | **Perceived stress –**<br>**adjusted model\*\*** |
| **Baseline**<br>N = 182 |  | *Reference* | *Reference* |
| **3 months**<br>N = 174 | *Estimate (95% CI), p* | *0.32 (-0.7 to 1.4)*<br>*P = 0.56* | *0.31 (-0.5 to 1.6)*<br>*P = 0.31* |
| **12 months**<br>N = 166 | *Estimate (95% CI), p* | *0.32 (-0.8 to 1.4)*<br>*P = 0.56* | *0.29 (-0.5 to 1.7)*<br>*P = 0.29* |
| *Changes in the FHT group* |  |  |  |
|  |  | **Perceived stress –**<br>**raw model\*** | **Perceived stress –**<br>**adjusted model\*\*** |
| **Baseline**<br>N = 144 |  | *Reference* | *Reference* |
| **3 months**<br>N = 129 | *Estimate (95% CI), p* | *-0.73 (-1.9 to 0.4)*<br>*P = 0.20* | *0.53 (-1.7 to 0.6)*<br>*P = 0.37* |
| **12 months**<br>N = 114 | *Estimate (95% CI), p* | *0.26 (-0.9 to 1.5)*<br>*P = 0.67* | *0.69 (-0.5 to 1.9)*<br>*P = 0.27* |
| *Changes in the FHT group compared to the MHT group* |  |  |  |
|  |  | **Perceived stress –**<br>**raw model\*** | **Perceived stress –**<br>**adjusted model\*\*** |
| **Baseline** |  | *Reference* | *Reference* |
| **3 months** | *Estimate (95% CI), p* | *-1.03 (-2.6 to 0.5)*<br>*P = 0.20* | *-1.0 (-2.6 to 0.6)*<br>*P = 0.21* |
| **12 months** | *Estimate (95% CI), p* | *0.00 (-1.6 to 1.6)*<br>*P = 0.99* | *0.25 (-1.4 to 1.9)*<br>*P = 0.76* |

\*Participants nested within countries.
\*\*Participants nested within countries, and adjusted for age and psychotropic medication use.

### Sensitivity analyses

Among 154 participants who provided data at all measurement time points (93 starting MHT and 61 starting FHT), stress levels did not significantly change after GAHT (Supplementary Materials 3).

Excluding FHT participants using estrogens only yielded similar results (Supplementary Materials 4). Adjusting for cycle regulation use (MHT: 36 [20%] at baseline, 32 [19%] at 3 months, 26 [16%] at 12 months) did not alter outcomes (Supplementary Materials 5).

### Post-hoc analyses

Initial analyses showed higher perceived stress scores in Belgium and Israel than in the Netherlands. Supplementary materials 2 shows that country was a significant predictor in model 3. To further explore these differences, post-hoc analyses were conducted, adjusting for demographic differences across countries (age, psychotropic medication use, alcohol use, smoking, and cycle regulation use).

Supplementary materials 6 presents the demographic characteristics by country. At all time points, participants in Belgium and Israel had higher stress levels than participants in the Netherlands. Table 5 presents PSS scores by country, and Figure 3 visualizes longitudinal trajectories in PSS scores per country.

**Figure 3.**
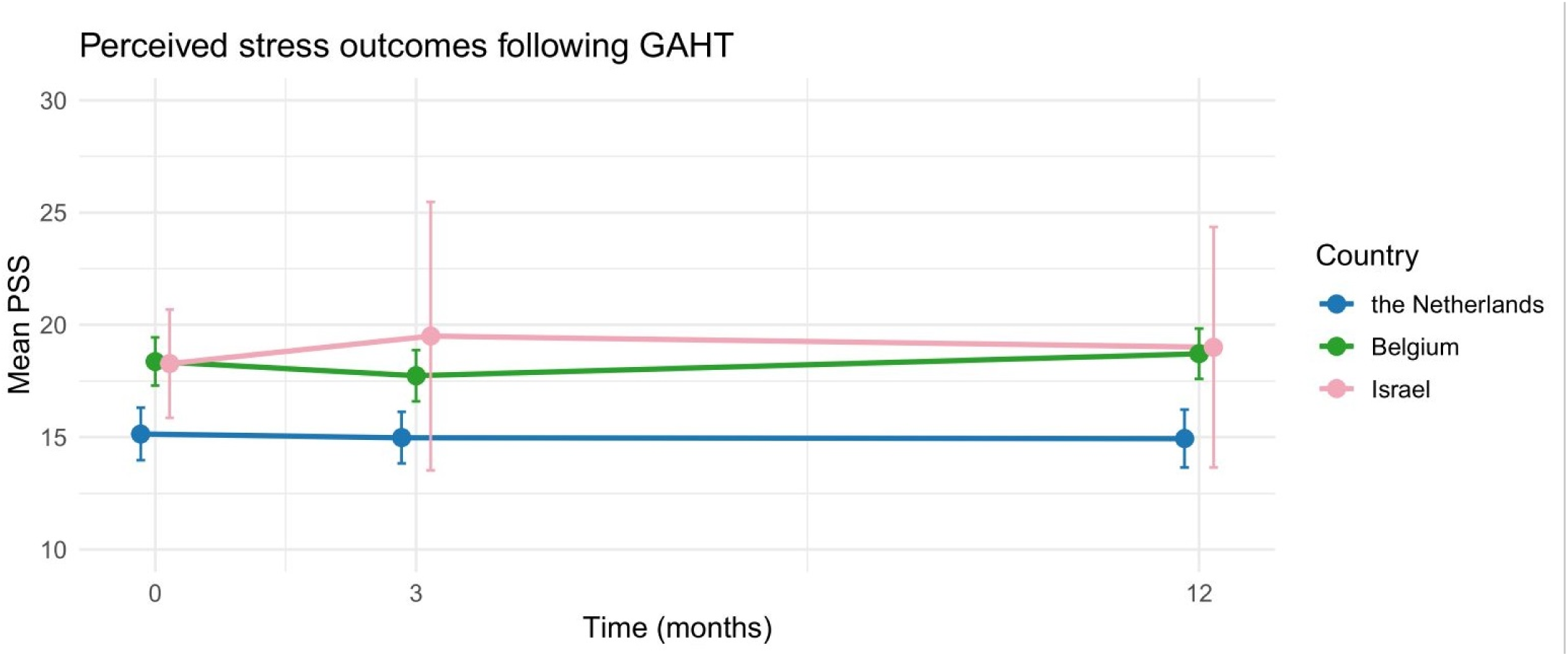
Mean PSS scores by country before GAHT, and after 3 and 12 months. The plot shows unadjusted group means per country at each time point, with error bars representing 95% confidence intervals around the observed means. Blue: the Netherlands, green: Belgium, pink: Israel.

**Table 5.** Perceived stress outcomes in each country.

| **Table 5. Perceived stress outcomes in each country** |  |  |  |
| --- | --- | --- | --- |
| **Baseline** |  |  |  |
|  |  | **Raw model** | **Adjusted model\*** |
| **The Netherlands**<br>N = 167 | *Mean (SD)* | 15.1 (7.7)<br>*Reference* | *Reference* |
| **Belgium**<br>N = 130 | *Mean (SD)*<br>*Estimate (95% CI), p* | 18.4 (6.2)<br>3.2 (1.6 to 4.8)<br>*P < 0.001* | 3.1 (1.4 to 4.8)<br>*P < 0.001* |
| **Israel**<br>N = 29 | *Mean (SD)*<br>*Estimate (95% CI), p* | 18.3 (6.3)<br>3.1 (0.6 to 5.9)<br>*P = 0.027* | 2.8 (-0.2 to 5.7)<br>*P = 0.065* |
| **3 months** |  |  |  |
|  |  | **Raw model** | **Adjusted model\*** |
| **The Netherlands**<br>N = 160 | *Mean (SD)* | 15.0 (7.3)<br>*Reference* | *Reference* |
| **Belgium**<br>N = 133 | *Mean (SD)*<br>*Estimate (95% CI), p* | 17.7 (6.6)<br>2.8 (1.1 to 4.4)<br>*P < 0.001* | 3.1 (0.9 to 4.3)<br>*P < 0.001* |
| **Israel**<br>N = 10 | *Mean (SD)*<br>*Estimate (95% CI), p* | 19.5 (8.4)<br>4.5 (0.0 to 9.0)<br>*P = 0.050* | 5.1 (0.2 to 9.1)<br>*P = 0.030* |
| **12 months** |  |  |  |
|  |  | **Raw model** | **Adjusted model\*** |
| **The Netherlands**<br>N = 139 | *Mean (SD)* | 14.9 (7.7)<br>*Reference* | *Reference* |
| **Belgium**<br>N = 137 | *Mean (SD)*<br>*Estimate (95% CI), p* | 18.7 (6.6)<br>3.8 (2.1 to 5.5)<br>*P < 0.001* | 3.4 (1.5 to 5.3)<br>*P < 0.001* |
| **Israel**<br>N = 4 | *Mean (SD)*<br>*Estimate (95% CI), p* | 19.0 (3.4)<br>4.1 (-3.1 to 11.2) | 6.1 (-1.7 to 13.9) |
| | | $P = 0.26$ | $P = 0.13$ |
| *Adjusted for age, psychotropic medication use, alcohol use, smoking, and cycle regulation use. |  |  |  |

## Discussion

This international multicenter prospective study assessed changes in perceived stress after 3 and 12 months of GAHT and explored differences between FHT and MHT. Before GAHT, stress levels did not significantly differ between the MHT and FHT groups. After 3 and 12 months of GAHT, no significant changes were observed compared to baseline, nor were there significant differences between FHT and MHT in these changes.

Our findings align with Ceolin et al. (2024), who also found no significant changes in perceived stress after 12 months of GAHT. This contrasts with studies reporting significant reductions in depressive symptoms and psychological distress (Chen et al., 2023; Colizzi et al., 2013; Doyle et al., 2023), and improved quality of life (Baker et al., 2021; van Leerdam et al., 2023). However, these improvements may not necessarily translate to similar changes for perceived stress, as perceived stress may encompass a broader evaluation of life circumstances. Perceived stress may also be influenced by persistent external gender minority stressors such as societal stigma or discrimination (Mezza et al., 2024; Singh et al., 2023), which may be unlikely to resolve within the first year of GAHT. Additionally, it is important to note that studies investigating psychological wellbeing after GAHT report mixed findings, with for example Morssinkhof et al. (2024) reporting worsening in low mood after 12 months of FHT.

Our findings contrast with those of Colizzi et al. (2013), who reported significant improvements in perceived stress after 12 months of GAHT. However, differences in baseline stress levels may help explain this discrepancy: Colizzi et al. (2013) observed an average of 27.7 (SD: 6.1), notably higher than in our study (FHT: 16.3, MHT: 17.1). Since a PSS score of 14-26 indicates moderate stress (Cohen et al., 1983), lower baseline levels in our cohort may explain the absence of significant change. The timing of the PSS measurements were also different in Colizzi et al. (2013) compared to our study: crucially, participants in Colizzi et al. (2013) were awaiting GAHT approval, a phase often associated with heightened uncertainty and stress about access to GAHT, while most in our study had already received approval, potentially lowering stress at baseline. Additionally, generational and cohort differences may play a role, as access to gender-affirming care and societal attitudes have evolved over the past decade. Recent studies report similar baseline PSS scores: Collet et al. (2023) found a median of 18.0 (IQR: 13.0-23.0), and Ceolin et al. (2024) reported means of 19.1 (SD: 6.4) for MHT and 18.2 (SD: 5.7) for FHT. This alignment in baseline stress levels likely contributes to the similar conclusions of no significant changes after GAHT across these studies.

We hypothesized significant differences in perceived stress after 3 and 12 months of FHT compared to MHT. For example, Foster Skewis et al. (2021) found that MHT improved emotional well-being and social functioning, whereas FHT did not. Additionally, FHT has been associated with greater increases in variability in negative affect (Morssinkhof et al., 2025) and worsening of low mood (Morssinkhof et al., 2024), which may contribute to increased stress perceptions alongside lower psychological well-being (Houben et al., 2015). However, no significant differences emerged between FHT and MHT, consistent with Colizzi et al. (2013).

Our hypothesis was partially based on biological factors, specifically changes in sex hormone levels. As reported in Supplementary materials 1, FHT resulted in large estradiol increases and testosterone decreases and MHT resulted in testosterone increases. While relationships between large changes in testosterone and estradiol levels and perceived stress remain unclear, insights can be drawn from research on fluctuations and stable sex hormone levels in cisgender women. Limited studies suggest that fluctuating estradiol levels, such as those occurring during the menstrual cycle or perimenopause, may amplify perceived stress more than stable increases or decreases in estradiol (Gordon et al., 2016; Gordon et al., 2019; Handy et al., 2022). Conversely, stable estradiol levels, as seen in studies on oral contraceptive use in premenopausal women or transdermal estrogen therapy in perimenopausal women, are generally associated with reduced perceived stress and greater mood stability (Gordon et al., 2018; Hamstra et al., 2017; Taggart et al., 2018). Although FHT-induced estradiol changes are not entirely stable, they appear less fluctuating than those observed during the menstrual cycle or perimenopause. This may explain why FHT does not significantly increase or decrease perceived stress levels. Similarly, MHT showed no significant association with changes in perceived stress. In our study, participants use different forms of testosterone, some of which induce more stable testosterone levels (i.e., testosterone gel) and others induce more fluctuating testosterone levels (i.e., short-acting testosterone injections). Unfortunately, research on the relationship between testosterone fluctuations and perceived stress is sparse, but future studies could examine the role of testosterone fluctuations in mood and perceived stress. Altogether, it is important to note that direct comparisons between our findings and those from other hormone intervention studies are challenging due to differences in hormonal composition, dosages, and clinical indications.

Psychosocial factors also played a role in our hypothesis, as individuals starting FHT may experience heightened gender minority stress (Poquiz et al., 2021). However, this was not reflected in our findings. Nonetheless, post-hoc analyses showed that participants from Belgium and Israel reported higher stress levels than those in the Netherlands before and after GAHT. Although the Netherlands and Belgium have a similar highly centralized gender-affirming healthcare structure (Kiely et al., 2024), societal acceptance of LGBTI+ individuals and gender diversity varies. The Netherlands ranks 2nd globally in social acceptance of LGBTI+ individuals, while Belgium ranks 15th, and Israel ranks 44th (Williams Institute, 2021), suggesting societal stressors may contribute to these differences.

Furthermore, based on visual inspection, the levels of perceived stress among participants in Belgium and Israel did not reduce to levels similar to those in the Netherlands after GAHT. Persistent impact of these external stressors, despite using GAHT, is a potential explanation for the absence of improvement in perceived stress. Moreover, as reported in Supplementary materials 6, psychotropic medication use was higher in Belgium (24%) and Israel (44%) than in the Netherlands (9%). These differences may reflect mental health disparities, potentially influenced by factors such as healthcare access, social acceptance or economic conditions. These factors may influence how participants experience and cope with stress.

The current study has several strengths. To begin, this is the first international multicenter study to examine differences in perceived stress among individuals starting MHT and FHT. The design allowed us to explore changes in stress alongside significant shifts in testosterone and estradiol levels. Second, the validated 10-item PSS was used, which effectively captures subjective stress. We believe that self-reported stress is a clinically more relevant measure than stress hormone levels, as they do not always align (Gidlow et al., 2016; Prado-Gascó et al., 2019; Schnall et al., 2024) and subjective experiences of stress often have a substantial impact on health outcomes (Gao et al., 2022; Russell & Lightman, 2019). Third, our linear mixed model accounted for individual- and country-level variability, offering more accurate estimates than treating country as a covariate (Moerbeek, 2004; Snijders & Bosker, 1999). Finally, post-hoc analyses across countries highlighted the role of societal and healthcare differences in shaping stress levels, enhancing the generalizability and highlighting the relevance of contextual factors in stress outcomes during GAHT.

Nevertheless, several limitations must be acknowledged. First, we lacked contextual information on relevant psychosocial factors. Perceived stress is influenced by various elements, and it remains unclear which – objective stressors, subjective perceptions, or coping mechanisms – played a role. Individuals with adequate coping resources may not report high stress despite significant stressor exposure, while others with fewer resources may find the same stressors overwhelming. Moreover, participants’ expectations of GAHT may influence perceived stress, with unmet needs (e.g., for social affirmation or additional interventions) or unrealistic expectations potentially contributing to unchanging stress levels. A review by Mezza et al. (2024) noted that distal (e.g., societal discrimination) and proximal stressors (e.g., internalized transphobia) are strongly linked to poor mental health outcomes. A recent daily diary study also found that microaggressions were associated with higher depressive and anxiety symptoms and lower self-esteem (Doyle et al., 2024). While our post-hoc findings showed that participants in Belgium and Israel reported significantly higher stress than those in the Netherlands, reflecting societal acceptance rankings for transgender individuals (Williams Institute, 2021), the lack of data on stressors and coping mechanisms limits our understanding of their impact on perceived stress during GAHT. Second, psychiatric comorbidities were not assessed in our cohort. However, the prevalence of psychotropic medication use (19% of those starting MHT and 16% of those starting FHT) was similar to that in the general adult population (Terlizzi & Norris, 2021). Third, participants from only three European countries were included, and post-hoc analyses showed significant differences in reported stress across countries. While this multi-country inclusion offers insights into social factors influencing perceived stress during GAHT, the results cannot be generalized to other countries, including those with higher societal stigma against transgender individuals. Lastly, our study used a 12-month follow-up period, but stress outcomes may take longer to manifest as gender minority stressors may persist. Longer follow-up periods might reveal different trends (Chen et al., 2023).

In conclusion, while previous studies showed significant improvements in psychological distress and quality of life after GAHT, our findings suggest these may not directly translate to perceived stress. The lack of significant differences between FHT and MHT indicates that sex hormone changes may not directly influence perceived stress in this context. A key limitation is the absence of contextual data on gender minority stressors and coping mechanisms. Future research should explore their role in shaping perceived stress during GAHT, offering insights into the pathways underlying potential stress improvements.

## Data Availability

All data produced in the present study are available upon reasonable request to the authors.

## CRediT statement

**Marijn Schipper:** Conceptualization, Methodology, Formal analysis, Data Curation, Writing – Original Draft. **Margot W. L. Morssinkhof:** Conceptualization, Methodology, Formal analysis, Investigation, Writing – Review & Editing. **Baudewijntje P. C. Kreukels:** Conceptualization, Writing – Review & Editing. **Guy T’Sjoen:** Resources, Investigation, Writing – Review & Editing. **Alessandra D. Fisher:** Resources, Investigation. **Yona Greenman:** Resources, Investigation, Writing – Review & Editing. **Karin van der Tuuk:** Resources, Investigation. **Martin den Heijer:** Resources, Investigation, Writing – Review & Editing. **David Matthew Doyle:** Conceptualization, Formal Analysis, Writing – Review & Editing, Supervision, Funding Acquisition. **Birit F. P. Broekman:** Conceptualization, Investigation, Writing – Review & Editing, Supervision, Funding Acquisition.

All authors agree to be accountable for all aspects of the work.

## Acknowledgements

We thank the participants of the ENIGI and RESTED studies, whose involvement was crucial to this research.

## Disclosures

The authors have no competing interests to disclose.

The RESTED study was funded by ZonMW/Netherlands Organisation for Health Research and Development under a Veni grant awarded to BB (91619085, 2018) and by the European Research Council under an ERC Starting Grant awarded to DD (ERC-StG 101042028). Views and opinions expressed are however those of the authors only and do not necessarily reflect those of the European Union or the European Research Council. Neither the European Union nor the granting authority can be held responsible for them.

Data in the current study are available upon reasonable request to the authors.

## Supplementary materials

### 1. Sex hormone levels

Supplementary table 1 reports sex hormone levels in serum at start and after 3 and 12 months of GAHT. The timing of hormone measurements was not adjusted for the type or timing of GAHT, the menstrual cycle, or cycle regulation use. Since endogenous sex hormone levels fluctuate diurnally and monthly, and exogenous hormone use induces fluctuations as well, the hormone measurements are considered suitable for assessing therapy adherence rather than causal effects.

**Supplementary table 1.** Sex hormone levels in serum at start of GAHT and after 3 and 12 months of GAHT.

|  |  |  |  |
| --- | --- | --- | --- |
|  |  | **MHT group** | **FHT group** |
| **Testosterone level at start of GAHT\*** (mean, SD) | *No CR use* | 1.07 nmol/L (1.13)<br>N = 135 | 16.48 nmol/L (6.4)<br>N = 130 |
|  | *CR use* | 0.91 nmol/L (1.15)<br>N = 33 |  |
| **Testosterone level after 3 months of GAHT\*** (mean, SD) | *No CR use* | 18.57 nmol/L (16.0)<br>N = 110 | 0.96 nmol/L (2.22)<br>N = 109 |
|  | *CR use* | 18.65 nmol/L (17.3)<br>N = 28 |  |
| **Testosterone level after 12 months of GAHT\*** (mean, SD) | *No CR use* | 19.89 nmol/L (11.7)<br>N = 89 | 1.30 nmol/L (2.6)<br>N = 94 |
|  | *CR use* | 14.63 nmol/L (9.7)<br>N = 21 |  |
| **Estradiol level at start of GAHT\*\*** (mean, SD) | *No CR use* | 235.01 pmol/L (251.5)<br>N = 127 | 77.18 pmol/L (29.8)<br>N = 126 |
|  | *CR use* | 136.01 pmol/L (179.9)<br>N = 31 |  |
| **Estradiol level after 3 months of GAHT\*\*** (mean, SD) | *No CR use* | 164.55 pmol/L (121.6)<br>N = 103 | 332.74 pmol/L (353.0)<br>N = 107 |
|  | *CR use* | 130.74 pmol/L (76.9)<br>N = 28 |  |
| **Estradiol level after 12 months of GAHT\*\*** (mean, SD) | *No CR use* | 126.92 pmol/L (67.4)<br>N = 93 | 324.04 pmol/L (208.1)<br>N = 90 |
|  | *CR use* | 149.73 pmol/L (99.7)<br>N = 20 |  |
| CRuse = cycle regulation use |  |  |  |

### 2. Linear mixed model fits

Supplementary table 2 shows the linear mixed models fits. Nesting participants within countries (model 4) showed a better fit than adding country as a covariate (model 3).

**Supplementary table 2.**
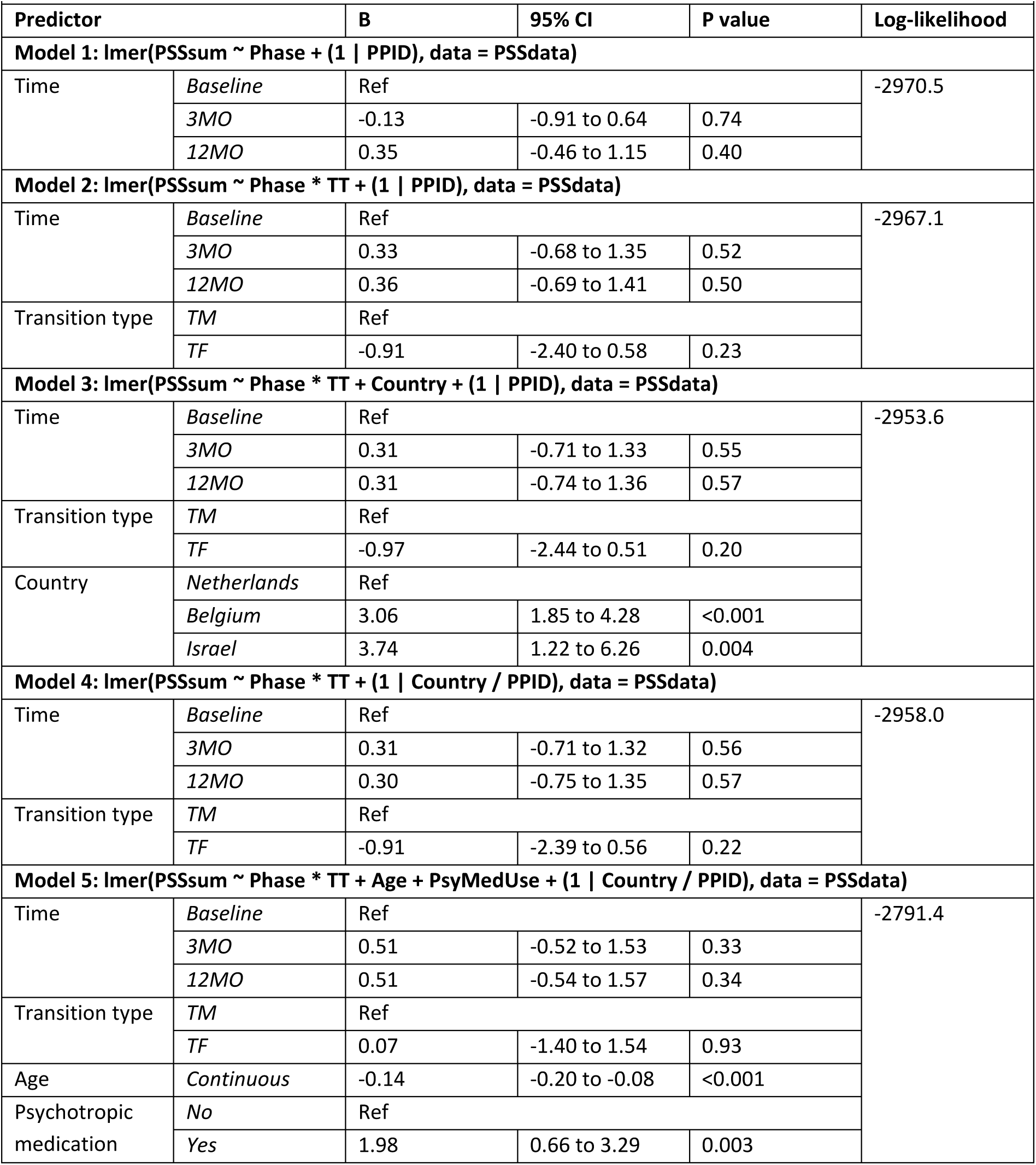
Linear mixed models.

### 3. Sensitivity analyses: Participants who contributed all measurement timepoints

The sample of participants who provided data to all measurement timepoints consisted of 154 persons, of whom 93 started MHT and 61 started FHT. Supplementary table 3 reports perceived stress outcomes at baseline. Perceived stress outcomes did not significantly differ between both groups before starting GAHT.

**Supplementary table 3.** Perceived stress outcomes before starting GAHT (baseline)

|  |  |  |  |  |
| --- | --- | --- | --- | --- |
|  |  | **Perceived stress – raw model** | **Perceived stress – nested model\*** | **Perceived stress – adjusted model\*\*** |
| **MHT group**<br>N = 182 | *Mean (SD)* | 16.96 (7.1) |  |  |
| **FHT group**<br>N = 144 | *Mean (SD)* | 15.28 (7.4) |  |  |
| **FHT group vs. MHT group** | *Estimate (95% CI), p* | -1.68 (-4.0 to 0.7)<br>P = 0.16 | -1.2 (-3.5 to 1.1)<br>P = 0.29 | -0.43 (-2.8 to 2.0)<br>P = 0.73 |
| *Participants nested within countries (i.e., the Netherlands, Belgium or Israel). |  |  |  |  |
| **Participants nested within countries, and adjusted for age and psychotropic medication use. |  |  |  |  |

Supplementary table 4 reports PSS outcomes after 3 and 12 months of GAHT. Perceived stress outcomes did not significantly change following 3 and 12 months of GAHT compared to baseline and no significant differences were observed between both groups.

**Supplementary table 4.** Perceived stress outcomes after 3 and 12 months of GAHT.

|  |  |  |  |  |
| --- | --- | --- | --- | --- |
| **Changes in the overall cohort** |  |  |  |  |
|  |  | **Perceived stress – raw model** | **Perceived stress – nested model\*** | **Perceived stress – adjusted model\*\*** |
| **Baseline**<br>N = 154 | *Mean (SD)*<br>*Reference* | 16.29 (7.2)<br>*Reference* | *Reference* | *Reference* |
| **3 months**<br>N = 154 | *Mean (SD)*<br>*Estimate (95% CI), p* | 16.05 (7.1)<br>-0.24 (-1.2 to 0.8)<br>P = 0.64 | -0.24 (-1.2 to 0.8)<br>P = 0.64 | -0.08 (-1.1 to 0.9)<br>P = 0.87 |
| **12 months**<br>N = 154 | *Mean (SD)*<br>*Estimate (95% CI), p* | 16.71 (7.6)<br>0.42 (-0.6 to 1.4)<br>P = 0.41 | 0.42 (-0.6 to 1.4)<br>P = 0.41 | 0.51 (-0.5 to 1.5)<br>P = 0.32 |
| **Changes in the MHT group** |  |  |  |  |
|  |  | **Perceived stress – raw model** | **Perceived stress – nested model\*** | **Perceived stress – adjusted model\*\*** |
| **Baseline**<br>N = 93 | *Mean (SD)*<br>*Reference* | 16.96 (7.1)<br>*Reference* | *Reference* | *Reference* |
| **3 months**<br>N = 93 | *Mean (SD)*<br>*Estimate (95% CI), p* | 16.95 (7.1)<br>-0.01 (-1.3 to 1.3)<br>P = 0.99 | -0.01 (-1.3 to 1.3)<br>P = 0.99 | 0.13 (-1.2 to 1.4)<br>P = 0.84 |
| **12 months**<br>N = 93 | *Mean (SD)*<br>*Estimate (95% CI), p* | 17.46 (7.9)<br>0.51 (-0.8 to 1.8)<br>P = 0.46 | 0.51 (-0.8 to 1.8)<br>P = 0.46 | 0.52 (-0.8 to 1.8)<br>P = 0.44 |

| Changes in the FHT group |  |  |  |  |
| --- | --- | --- | --- | --- |
|  |  | Perceived stress –<br>raw model | Perceived stress –<br>nested model* | Perceived stress –<br>adjusted model** |
| **Baseline**<br>N = 61 | Mean (SD)<br>Reference | 15.28 (7.4)<br>Reference | Reference | Reference |
| **3 months**<br>N = 61 | Mean (SD)<br>Estimate (95% CI), p | 14.69 (7.0)<br>-0.59 (-2.1 to 0.9)<br>P = 0.45 | -0.59 (-2.1 to 0.9)<br>P = 0.45 | -0.40 (-1.9 to 1.1)<br>P = 0.61 |
| **12 months**<br>N = 61 | Mean (SD)<br>Estimate (95% CI), p | 15.57 (7.0)<br>0.30 (-1.2 to 1.8)<br>P = 0.70 | 0.30 (-1.2 to 1.8)<br>P = 0.70 | 0.60 (-0.9 to 2.1)<br>P = 0.44 |
| Changes in the FHT group compared to the MHT group |  |  |  |  |
|  |  | Perceived stress –<br>raw model | Perceived stress –<br>nested model* | Perceived stress –<br>adjusted model** |
| **Baseline** | Reference | Reference | Reference | Reference |
| **3 months** | Estimate (95% CI), p | -0.58 (-2.6 to 1.5)<br>P = 0.58 | -0.58 (-2.6 to 1.5)<br>P = 0.58 | -0.71 (-2.7 to 1.3)<br>P = 0.49 |
| **12 months** | Estimate (95% CI), p | -0.21 (-2.3 to 1.8)<br>P = 0.84 | -0.21 (-2.3 to 1.8)<br>P = 0.84 | -0.08 (-2.1 to 1.9)<br>P = 0.94 |
| *Participants nested within countries. |  |  |  |  |
| **Participants nested within countries, and adjusted for age and psychotropic medication use. |  |  |  |  |

### 4. Sensitivity analyses: Only combined FHT

Supplementary table 5 reports sensitivity analyses excluding participants using only estrogens instead of a combination of estrogens and anti-androgens. These outcomes after 3 and 12 months of GAHT were similar to the results in the main paper.

**Supplementary table 5.** Perceived stress outcomes after 3 and 12 months of GAHT.

|  |  |  |  |
| --- | --- | --- | --- |
| **Changes in the FHT group** |  |  |  |
|  | **Perceived stress –**<br>**raw model** | **Perceived stress –**<br>**nested model\*** | **Perceived stress –**<br>**adjusted model\*\*** |
| **Baseline**<br>N = 138 | 16.14 (7.1)<br>*Reference* | *Reference* | *Reference* |
| **3 months**<br>N = 126 | 14.91 (6.9)<br>-0.92 (-2.1 to 0.2)<br>*P* = 0.12 | -0.91 (-2.1 to 0.2)<br>*P* = 0.12 | -0.71 (-1.9 to 0.5)<br>*P* = 0.24 |
| **12 months**<br>N = 113 | 16.22 (7.3)<br>0.22 (-1.0 to 1.4)<br>*P* = 0.73 | 0.14 (-1.1 to 1.3)<br>*P* = 0.82 | 0.57 (-0.7 to 1.8)<br>*P* = 0.36 |
| **Changes in the FHT group compared to the MHT group** |  |  |  |
|  | **Perceived stress –**<br>**raw model** | **Perceived stress –**<br>**nested model\*** | **Perceived stress –**<br>**adjusted model\*\*** |
| **Baseline** | *Reference* | *Reference* | *Reference* |
| **3 months** | -1.27 (-2.8 to 0.3)<br>*P* = 0.12 | -1.19 (-2.8 to 0.4)<br>*P* = 0.14 | -1.17 (-2.7 to 0.4)<br>*P* = 0.14 |
| **12 months** | -0.15 (-1.8 to 1.5)<br>*P* = 0.86 | -0.10 (-1.7 to 1.5)<br>*P* = 0.91 | 0.13 (-1.5 to 1.8)<br>*P* = 0.88 |
| *Participants nested within countries. |  |  |  |
| **Participants nested within countries, and adjusted for age and psychotropic medication use. |  |  |  |

### 5. Sensitivity analyses: Cycle regulation use

Supplementary table 6 reports cycle regulation use per timepoint. Among participants in the MHT group, cycle regulation was used at baseline by 36 (20%) participants, after 3 months of GAHT by 32 (19%) participants and after 12 months of GAHT by 26 (16%) participants.

**Supplementary table 6.** Number of participants using cycle regulation at baseline and after 3 and 12 months of MHT.

|  |  |  |
| --- | --- | --- |
|  | **No cycle regulation use** | **Cycle regulation use** |
| **Baseline** (n, %) | 141 (80%) | 36 (20%) |
| **3 months** (n, %) | 133 (81%) | 32 (19%) |
| **12 months** (n, %) | 134 (84%) | 26 (16%) |

Supplementary table 7 reports changes in perceived stress after adjusting for cycle regulation use among participants in the MHT group. The outcomes were similar to those observed without adjusting for cycle regulation.

**Supplementary table 7.** Perceived stress outcomes after 3 and 12 months of GAHT, after adjusting for cycle regulation use.

|  |  |
| --- | --- |
| **Changes in the MHT group** |  |
|  | **Adjusted model\*** |
| *CR use vs. no CR use* | 1.58 (-1.1 to 4.3)<br>*P* = 0.24 |
| *3 months of GAHT vs. baseline* | 0.13 (-1.2 to 1.4)<br>*P* = 0.85 |
| *12 months of GAHT vs. baseline* | 0.54 (-0.8 to 1.8)<br>*P* = 0.42 |
| *Participants were nested within countries, and the model was adjusted for age, psychotropic medication use and cycle regulation use. |  |

### 6. Post-hoc analyses: Demographic characteristics for each country

Supplementary table 8 reports the demographic characteristics of the participants per country. The median age of the participants in the Netherlands was 24.0 years (SD 20.8 to 29.2), of the participants in Belgium 22.7 years (SD 20.5 to 27.5), and of the participants in Israel 21.6 years (SD 20.0 to 25.5). Psychotropic medication was used by 9% of the participants in the Netherlands, whereas 24% of the participants in Belgium and 44% of participants in Israel used psychotropic medication. The participants in Belgium smoked cigarettes more often, as 26% of the participants in Belgium, 13% of the participants in Israel, and 11% of the participants in the Netherlands reported that they smoked cigarettes. Additionally, cycle regulation was used by 27% of the TM participants at baseline in the Netherlands, whereas 13% of the TM participants in Belgium and no TM participants in Israel used cycle regulation at baseline.

**Supplementary table 8.**
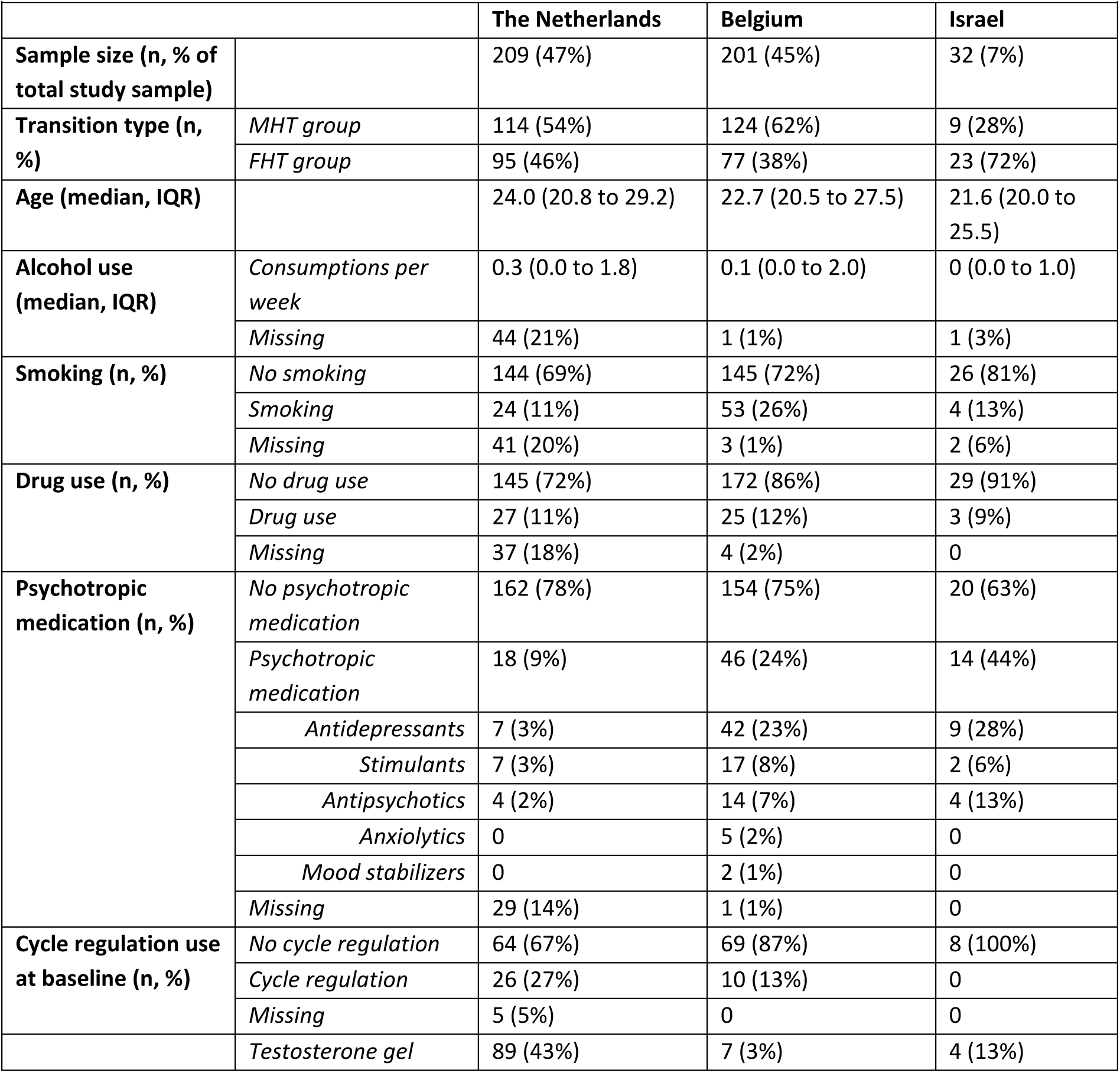
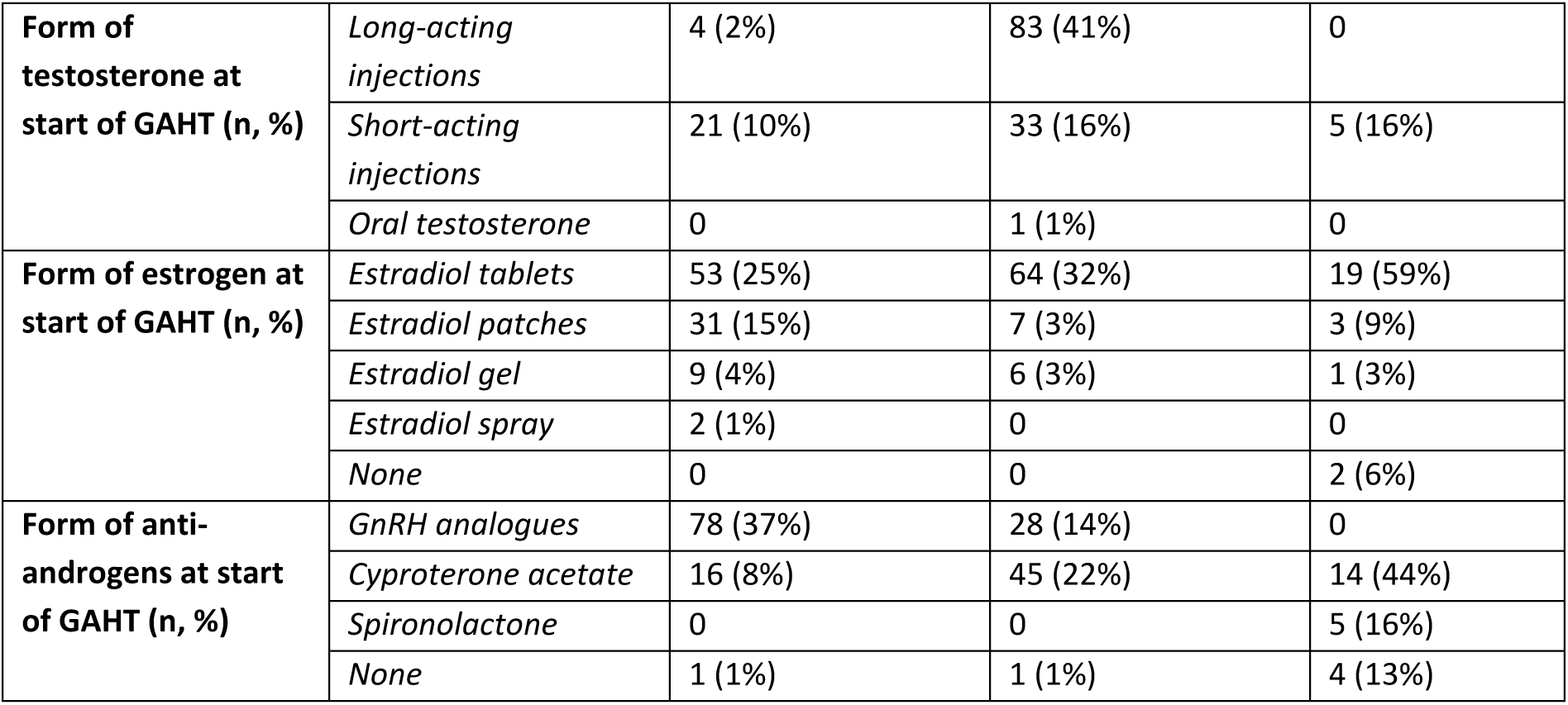
Demographic characteristics of the study participants in each country.

## References

American Psychological Association. (2022). *Stress in America* 2022. Retrieved on 2024-12-17, from https://www.apa.org/news/press/releases/stress/2022/concerned-future-inflation

Baker, K. E., Wilson, L. M., Sharma, R., Dukhanin, V., McArthur, K., & Robinson, K. A. (2021). Hormone Therapy, Mental Health, and Quality of Life Among Transgender People: A Systematic Review. J Endocr Soc, 5(4), bvab011.

Bennett, J. M., Rohleder, N., & Sturmberg, J. P. (2018). Biopsychosocial approach to understanding resilience: Stress habituation and where to intervene. J Eval Clin Pract, 24(6), 1339–1346.

Call, D. C., Challa, M., & Telingator, C. J. (2021). Providing Affirmative Care to Transgender and Gender Diverse Youth: Disparities, Interventions, and Outcomes. Curr Psychiatry Rep, 23(6), 33.

Ceolin, C., Scala, A., Scagnet, B., Citron, A., Vilona, F., De Rui, M., . . . Garolla, A. (2024). Body composition and perceived stress levels in transgender individuals after one year of gender affirming hormone therapy. Front Endocrinol (Lausanne), 15, 1496160.

Chen, D., Berona, J., Chan, Y. M., Ehrensaft, D., Garofalo, R., Hidalgo, M. A., . . . Olson-Kennedy, J. (2023). Psychosocial Functioning in Transgender Youth after 2 Years of Hormones. N Engl J Med, 388(3), 240–250.

Cohen, S., Kamarck, T., & Mermelstein, R. (1983). A global measure of perceived stress. J Health Soc Behav, 24(4), 385–396.

Colizzi, M., Costa, R., Pace, V., & Todarello, O. (2013). Hormonal treatment reduces psychobiological distress in gender identity disorder, independently of the attachment style. J Sex Med, 10(12), 3049–3058.

Collet, S., Kiyar, M., Martens, K., Vangeneugden, J., Simpson, V. G., Guillamon, A., . . . T’Sjoen, G. (2023). Gender minority stress in transgender people: a major role for social network. J Sex Med, 20(6), 905–917.

Dekker, M. J., Wierckx, K., Van Caenegem, E., Klaver, M., Kreukels, B. P., Elaut, E., . . . T’Sjoen, G. (2016). A European Network for the Investigation of Gender Incongruence: Endocrine Part. J Sex Med, 13(6), 994–999.

Doyle, D. M., Georgiev, D., Lewis, T. O. G., & Barreto, M. (2024). Frequency and mental health consequences of microaggressions experienced in the day-to-day lives of transgender and gender diverse people. International Journal of Transgender Health, 1-14.

Doyle, D. M., Lewis, T. O. G., & Barreto, M. (2023). A systematic review of psychosocial functioning changes after gender-affirming hormone therapy among transgender people. Nat Hum Behav, 7(8), 1320–1331.

Folkman, S. (2020). Stress: Appraisal and Coping. In M. D. Gellman (Red.), Encyclopedia of Behavioral Medicine (pp. 2177-2179). Cham: Springer International Publishing.

Foster Skewis, L., Bretherton, I., Leemaqz, S. Y., Zajac, J. D., & Cheung, A. S. (2021). Short-Term Effects of Gender-Affirming Hormone Therapy on Dysphoria and Quality of Life in Transgender Individuals: A Prospective Controlled Study. Front Endocrinol (Lausanne), 12, 717766.

Gao, Q., Xu, H., Zhang, C., Huang, D., Zhang, T., & Liu, T. (2022). Perceived stress and stress responses during COVID-19: The multiple mediating roles of coping style and resilience. PLoS One, 17(12), e0279071.

Gidlow, C. J., Randall, J., Gillman, J., Silk, S., & Jones, M. V. (2016). Hair cortisol and self-reported stress in healthy, working adults. Psychoneuroendocrinology, 63, 163–169.

Gordon, J. L., Eisenlohr-Moul, T. A., Rubinow, D. R., Schrubbe, L., & Girdler, S. S. (2016). Naturally Occurring Changes in Estradiol Concentrations in the Menopause Transition Predict Morning Cortisol and Negative Mood in Perimenopausal Depression. Clin Psychol Sci, 4(5), 919–935.

Gordon, J. L., Peltier, A., Grummisch, J. A., & Sykes Tottenham, L. (2019). Estradiol Fluctuation, Sensitivity to Stress, and Depressive Symptoms in the Menopause Transition: A Pilot Study. Front Psychol, 10, 1319.

Gordon, J. L., Rubinow, D. R., Eisenlohr-Moul, T. A., Xia, K., Schmidt, P. J., & Girdler, S. S. (2018). Efficacy of Transdermal Estradiol and Micronized Progesterone in the Prevention of Depressive Symptoms in the Menopause Transition: A Randomized Clinical Trial. JAMA Psychiatry, 75(2), 149–157.

Graves, B. S., Hall, M. E., Dias-Karch, C., Haischer, M. H., & Apter, C. (2021). Gender differences in perceived stress and coping among college students. PLoS One, 16(8), e0255634.

Hamstra, D. A., de Kloet, E. R., de Rover, M., & Van der Does, W. (2017). Oral contraceptives positively affect mood in healthy PMS-free women: A longitudinal study. Journal of Psychosomatic Research, 103, 119–126.

Handy, A. B., Greenfield, S. F., Yonkers, K. A., & Payne, L. A. (2022). Psychiatric Symptoms Across the Menstrual Cycle in Adult Women: A Comprehensive Review. Harv Rev Psychiatry, 30(2), 100–117.

Henze, G. I., Konzok, J., Kreuzpointner, L., Bärtl, C., Giglberger, M., Peter, H., . . . Wüst, S. (2021). Sex-specific interaction between cortisol and striato-limbic responses to psychosocial stress. Soc Cogn Affect Neurosci, 16(9), 972–984.

Houben, M., Van Den Noortgate, W., & Kuppens, P. (2015). The relation between short-term emotion dynamics and psychological well-being: A meta-analysis. Psychol Bull, 141(4), 901–930.

Hoyt, L. T., & Falconi, A. M. (2015). Puberty and perimenopause: reproductive transitions and their implications for women’s health. Soc Sci Med, 132, 103–112.

Keller, A., Litzelman, K., Wisk, L. E., Maddox, T., Cheng, E. R., Creswell, P. D., & Witt, W. P. (2012). Does the perception that stress affects health matter? The association with health and mortality. Health Psychol, 31(5), 677–684.

Kelly, M. M., Tyrka, A. R., Anderson, G. M., Price, L. H., & Carpenter, L. L. (2008). Sex differences in emotional and physiological responses to the Trier Social Stress Test. J Behav Ther Exp Psychiatry, 39(1), 87–98.

Kiely, E., Millet, N., Baron, A., Kreukels, B. P. C., & Doyle, D. M. (2024). Unequal geographies of gender-affirming care: A comparative typology of trans-specific healthcare systems across Europe. Soc Sci Med, 356, 117145.

Kirschbaum, C., Kudielka, B. M., Gaab, J., Schommer, N. C., & Hellhammer, D. H. (1999). Impact of gender, menstrual cycle phase, and oral contraceptives on the activity of the hypothalamus-pituitary-adrenal axis. Psychosom Med, 61(2), 154–162.

Kuehner, C. (2017). Why is depression more common among women than among men? Lancet Psychiatry, 4(2), 146–158.

Kuznetsova, A., Brockhoff, P. B., & Christensen, R. H. B. (2017). lmerTest package: tests in linear mixed effects models. Journal of statistical software, 82(13).

Lee, E. H. (2012). Review of the psychometric evidence of the perceived stress scale. Asian Nurs Res (Korean Soc Nurs Sci), 6(4), 121–127.

Leistner, C., & Menke, A. (2020). Hypothalamic-pituitary-adrenal axis and stress. Handb Clin Neurol, 175, 55–64.

Mezza, F., Mezzalira, S., Pizzo, R., Maldonato, N. M., Bochicchio, V., & Scandurra, C. (2024). Minority stress and mental health in European transgender and gender diverse people: A systematic review of quantitative studies. Clin Psychol Rev, 107, 102358.

Moerbeek, M. (2004). The Consequence of Ignoring a Level of Nesting in Multilevel Analysis. Multivariate Behav Res, 39(1), 129–149.

Morssinkhof, M. W. L., Schipper, M., Kreukels, B. P. C., van der Tuuk, K., den Heijer, M., van den Heuvel, O. A., . . . Broekman, B. F. P. (2025). Changes in affect variability after starting gender-affirming hormone therapy. Psychoneuroendocrinology, 175, 107408.

Morssinkhof, M. W. L., Wiepjes, C. M., van den Heuvel, O. A., Kreukels, B. P. C., van der Tuuk, K., T’Sjoen, G., . . . Broekman, B. F. P. (2024). Changes in depression symptom profile with gender-affirming hormone use in transgender persons. J Affect Disord, 348, 323–332.

Poquiz, J. L., Coyne, C. A., Garofalo, R., & Chen, D. (2021). Comparison of Gender Minority Stress and Resilience Among Transmasculine, Transfeminine, and Nonbinary Adolescents and Young Adults. J Adolesc Health, 68(3), 615–618.

Prado-Gascó, V., de la Barrera, U., Sancho-Castillo, S., de la Rubia-Ortí, J. E., & Montoya-Castilla, I. (2019). Perceived stress and reference ranges of hair cortisol in healthy adolescents. PLoS One, 14(4), e0214856.

R Development Core Team. (2010). R: A language and environment for statistical computing

Riecher-Rössler, A. (2017). Sex and gender differences in mental disorders. Lancet Psychiatry, 4(1), 8–9.

Russell, G., & Lightman, S. (2019). The human stress response. Nat Rev Endocrinol, 15(9), 525–534.

Salk, R. H., Hyde, J. S., & Abramson, L. Y. (2017). Gender differences in depression in representative national samples: Meta-analyses of diagnoses and symptoms. Psychol Bull, 143(8), 783–822.

Schnall, R., Liu, J., Cordoba, E., Brin, M., Garofalo, R., Kuhns, L. M., . . . Bendinskas, K. (2024). Differences in Self-Reported Stress Versus Hair and Nail Cortisol Among Adolescent and Young Adult Males. Nurs Res, 73(6), 442–449.

Seedat, S., Scott, K. M., Angermeyer, M. C., Berglund, P., Bromet, E. J., Brugha, T. S., . . . Kessler, R. C. (2009). Cross-national associations between gender and mental disorders in the World Health Organization World Mental Health Surveys. Arch Gen Psychiatry, 66(7), 785–795.

Singh, A., Dandona, A., Sharma, V., & Zaidi, S. Z. H. (2023). Minority Stress in Emotion Suppression and Mental Distress Among Sexual and Gender Minorities: A Systematic Review. Ann Neurosci, 30(1), 54–69.

Smith, W. C. (2021). Consequences of school closure on access to education: Lessons from the 2013-2016 Ebola pandemic. Int Rev Educ, 67(1-2), 53–78.

Snijders, T., & Bosker, R. (1999). Multilevel Analysis: An Introduction to Basic and Advanced Multilevel Modeling. http://lst-iiepiiep-unescoorg/cgi-bin/wwwi32exe/[in=epidoc1in]/?t2000=013777/(100).

Stephens, M. A., Mahon, P. B., McCaul, M. E., & Wand, G. S. (2016). Hypothalamic-pituitary-adrenal axis response to acute psychosocial stress: Effects of biological sex and circulating sex hormones. Psychoneuroendocrinology, 66, 47–55.

Stoica, T., Knight, L. K., Naaz, F., Patton, S. C., & Depue, B. E. (2021). Gender differences in functional connectivity during emotion regulation. Neuropsychologia, 156, 107829.

Taggart, T. C., Eaton, N. R., Keyes, K. M., Hammett, J. F., & Ulloa, E. C. (2018). Oral contraceptive use is associated with greater mood stability and higher relationship satisfaction. *Neurology*, Psychiatry and Brain Research, 30, 154–162.

Terlizzi, E. P., & Norris, T. (2021). Mental Health Treatment Among Adults: United States, 2020. NCHS Data Brief, (419), 1-8.

Thorsén, F., Antonson, C., Palmér, K., Berg, R., Sundquist, J., & Sundquist, K. (2022). Associations between perceived stress and health outcomes in adolescents. Child Adolesc Psychiatry Ment Health, 16(1), 75.

Turban, J. L., King, D., Kobe, J., Reisner, S. L., & Keuroghlian, A. S. (2022). Access to gender-affirming hormones during adolescence and mental health outcomes among transgender adults. PLoS One, 17(1), e0261039.

van Leerdam, T. R., Zajac, J. D., & Cheung, A. S. (2023). The Effect of Gender-Affirming Hormones on Gender Dysphoria, Quality of Life, and Psychological Functioning in Transgender Individuals: A Systematic Review. Transgend Health, 8(1), 6–21.

Williams Institute. (2021). Social Acceptance of LGBTI People in 175 Countries and Locations. Retrieved on 2024-12-17, from https://williamsinstitute.law.ucla.edu/publications/global-acceptance-index-lgbt/

Woods, N. F., Carr, M. C., Tao, E. Y., Taylor, H. J., & Mitchell, E. S. (2006). Increased urinary cortisol levels during the menopausal transition. Menopause, 13(2), 212–221.

Yalcin-Siedentopf, N., Pichler, T., Welte, A. S., Hoertnagl, C. M., Klasen, C. C., Kemmler, G., . . . Hofer, A. (2021). Sex matters: stress perception and the relevance of resilience and perceived social support in emerging adults. Arch Womens Ment Health, 24(3), 403–411.

Yılmaz Koğar, E., & Koğar, H. (2024). A systematic review and meta-analytic confirmatory factor analysis of the perceived stress scale (PSS-10 and PSS-14). Stress Health, 40(1), e3285.

